# Prediction-powered Inference for Clinical Trials: application to linear covariate adjustment

**DOI:** 10.1101/2025.01.15.25320578

**Authors:** Pierre-Emmanuel Poulet, Maylis Tran, Sophie Tezenas du Montcel, Bruno Dubois, Stanley Durrleman, Bruno Jedynak, the Alzheimers Disease Neuroimaging Initiative

## Abstract

Prediction-powered inference (PPI) [1] and its subsequent development called PPI++ [2] provide a novel approach to standard statistical estimation, leveraging machine learning systems, to enhance unlabeled data with predictions. We use this paradigm in clinical trials. The predictions are provided by disease progression models, providing prognostic scores for all the participants as a function of baseline covariates. The proposed method would empower clinical trials by providing untreated digital twins of the treated patients while remaining statistically valid. The potential implications of this new estimator of the treatment effect in a two-arm randomized clinical trial (RCT) are manifold. First, it leads to an overall reduction of the sample size required to reach the same power as a standard RCT. Secondly, it advocates for an imbalance of controls and treated patients, requiring fewer controls to achieve the same power. Finally, this technique directly transfers any disease prediction model trained on large cohorts to practical and scientifically valid use. In this paper, we demonstrate the theoretical properties of this estimator and illustrate them through simulations. We show that it is asymptotically unbiased for the Average Treatment Effect and derive an explicit formula for its variance. We then compare this estimator to a regression-based linear covariate adjustment method. An application to an Alzheimer’s disease clinical trial showcases the potential to reduce the sample size.

## 1 Introduction

Clinical trials are essential to demonstrate the benefits of a new treatment. Evaluating whether an intervention affects a measure of disease severity is one of the focus of causal inference [3]. Randomized clinical trials (RCT) provide a gold standard in this field. A key component in designing an RCT is estimating the treatment effect correctly and providing an uncertainty measure to perform a statistical test.

A primary lever to decrease uncertainty in a clinical trial’s outcome is to increase the sample size. However, recruiting more subjects has a cost, both financial and ethical, since we cannot provide the treatment to every participant. Moreover, enrolling enough patients for a rare disease in a short time window can be extremely challenging.

Some approaches have been proposed to use additional information from patients in other cohorts, like the placebo arm of another RCT or patients in observational cohorts. Among such methods, we can cite Matching Adjusted Indirect Comparisons (MAIC) [4], which adjusts for covariates distribution shifts between the compared cohort and the clinical trial of interest, or Bayesian Dynamical Borrowing, which uses data from the previous study under the form of a Bayesian prior [5]. They lead to increased power in clinical trials. However, these techniques usually lack control over the type I error rate.

Simultaneously, machine-learning techniques have been developed in almost every field of medicine. Specifically, we have witnessed a sharp increase in disease progression models (DPM) performance over the last few years. Whether they are event-based models such as SuStaIn [6], non-linear mixed effect models such as Disease Course Mapping [7] or dynamical models based on differential equations [8, 9], these models have shown great potential in predicting the evolution of a disease over time. Our approach will thus rely on such systems to learn the disease progression patterns from historical data. They can then help select or stratify patients for a clinical trial [10] and improve clinical trial efficacy [11]. In our case, we will use the trained models to predict the evolution of the patients in the clinical trial from their baseline information (and, when available, pre-inclusion information). This will yield a prognostic score that will be included in our analysis. This is akin to the digital twins’ methods [12].

By accounting for variability associated with prognostic factors, the precision of the estimated treatment effect can be improved, leading to narrower confidence intervals. Prior developments have been made to include prognostic factors in the statistical analysis of an RCT, among which the prognostic covariate adjustment method [13] which has been approved by regulatory agencies [14]. Even though this method is quite recent, the idea to adjust for prognostic scores or other composite scores is not new [15]. Our approach builds on the recent concept of prediction-powered inference (PPI) [1], and its subsequent extension PPI++ [2], a method which bridges machine learning prediction systems and statistical estimators by enhancing the latter. More specifically, the method guarantees a decrease in the variance of the estimators, meaning narrower confidence intervals and potential sample size reduction. An application to medical data was proposed in [16]. We extend this framework to RCT with disease progression models as prognostic factors.

Even though prognostic factor adjustment is essential to improve the validity and efficacy of RCTs by increasing their power or reducing sample size, they can be prone to overfitting when too many factors are included. Our approach relies on a single prognostic score, also denoted a super-covariate in [17], which, on the downside, requires this score to be accurate. Finally, every part of the method is pre-specifiable for an RCT analysis and can be used to determine the sample size needed at the design stage. Our contributions in this paper are the following: we propose adapting the prediction-powered estimators to clinical trials and derive their mathematical properties. We compare to ANCOVA [13], a linear covariate adjustment method. We perform basic simulations with small samples and unbalanced designs to contrast the different methods. We then perform a re-analysis of a randomized clinical trial for Alzheimer’s disease. First, using simulations, and then using the actual trial data. In each case, we contrast different estimators of the average treatment effect: a reference estimator, or baseline, PPI, and ANCOVA.

## 2 Method

Let us introduce our notations. Let *N* be the total number of patients (or participants) in the clinical trial. The patients, denoted by index *i* ∈ [1, *N*], are randomly assigned to the treated arm and the control arm. We use the words “treatment and control” generically while acknowledging that the control arm is often associated with a default treatment. For simplicity, we do not consider covariate-adaptive randomization in our analysis, such as stratified permuted block design [18] or Pocock-Simon’s minimization [19]. Without loss of generality, and for convenience of notation, we assume the first *m* patients to be in the treated arm while the *n* = *N* − *m* others are controls or placebo. The values of *m* and *n* can be adjusted in the trial’s design, allowing for an imbalance.

We denote (*X*_*i*_)_1≤*i*≤*N*_ ∈ 𝒳^*N*^ the baseline variables of the patients, (*Y*_*i*_)_1≤*i*≤*N*_ ∈ ℝ^*N*^ the outcome of the clinical trial, and (*T*_*i*_)_1≤*i*≤*N*_ ∈ {0, 1}^*N*^ the treatment variable, such that for 1 ≤ *i* ≤ *m, T*_*i*_ = 1 and for *m* + 1 ≤ *i* ≤ *N, T*_*i*_ = 0. We will often use 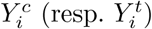 to denote 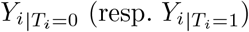. We assume throughout that given the randomization, the patients’ random variables are independent samples from a common distribution (*X*_*i*_, *Y*_*i*_ |*T*_*i*_) ∼𝒟. Adequate randomization ensures that *X*_*i*_ ⊥ *T*_*i*_.

The average treatment effect is our estimand, which is defined as follows:

### Definition 1

(ATE). *Using previously introduced notations, let the average treatment effect:*

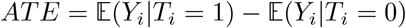

It is the difference between *θ*_1_ = 𝔼 (*Y*_*i*_|*T*_*i*_ = 1) and *θ*_0_ = 𝔼 (*Y*_*i*_|*T*_*i*_ = 0).

We assume that we are given a prediction function *f* : *X* ∈ 𝒳↦*f* (*X*) ∈ ℝ. In theory, this function can be any black-box predictor; in practice, it is designed to work best with a prognostic score such that *f* (*X*_*i*_) is highly correlated with *Y* ^*c*^. A special case of interest is when *f* is a machine-learning predictor. In this case, we require that *f* be trained with data that is disjoint from that of the trial.

We also introduce notations for the variance and covariance of the variables mentioned previously: 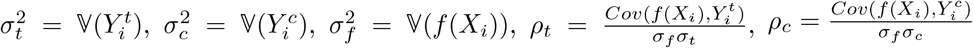.

### 2.1 Reference estimator

The simplest estimator for the average treatment effect, which we will call the reference estimator, is the following:

#### Definition 2

(Reference estimator). *Let the reference estimator for ATE:*

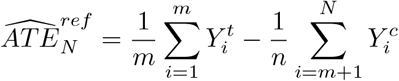

We often neglect the subscript *N* in the following to simplify notations. The Central Limit Theorem provides the proof of the next proposition:

#### Proposition 1.

*Assuming samples* (*Y*_*i*_, *X*_*i*_, *T*_*i*_)_*i*_ *are i*.*i*.*d, then:*

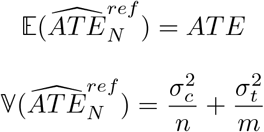

*and the estimator is asymptotically normal:*

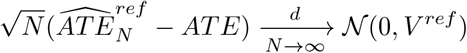

*where* 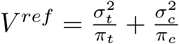 *with* 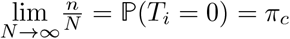 *and* 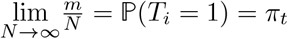.

From the variance formula we can derive an optimal partition between controls and treated patients: 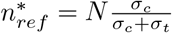 which yields 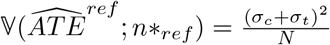. Designing a trial with this optimal partition implies that one has access to approximate values for the standard deviations of the outcome in the control arm and the treated arm, respectively *σ*_*c*_ and *σ*_*t*_. It can be done using a pilot study or a previous clinical trial. However, it is often assumed that *σ*_*t*_ = *σ*_*c*_ for simplicity, alleviating the estimation burden as the variance of the outcome under no intervention is often easier to estimate, for instance, using an observational cohort. Note that the reference estimator is seldom used when covariates are available.

### 2.2 Prediction-powered estimator

Prediction-powered inference (PPI) [2] relied on the intuition that it might be better to estimate *θ*_0_ from predictions (*f* (*X*_*i*_))_1≤*i*≤*N*_ rather than relying on the control observations (*Y*_*i*_)_*m<i*≤*N*_. However 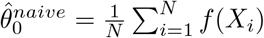 has no guarantee to be unbiased. Therefore a correction to the predictions is obtained by computing the empirical bias between control patients and their prediction: 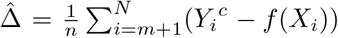, a term which is called the *rectifier*. Combining with the empirical mean for the estimator of *θ*_1_, we get:

#### Definition 3

(PPI estimator). *Using previous notations, let the prediction-powered inference estimator:*

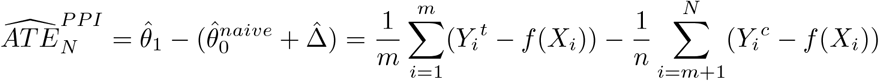

Using the CLT, we obtain:

#### Proposition 2.

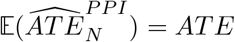

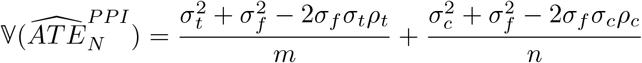

*and the estimator is asymptotically normal:*

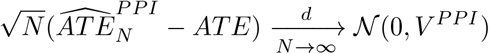

*where* 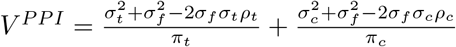

Note firstly that the prognostic models need not be unbiased. It is rectified by the observation gathered at the end of the trial. This is the most salient feature of PPI. Secondly, compared with the variance of the reference estimator, see proposition 1, the variance of the PPI estimator depends on the quality of the prediction function *f* through the terms *ρ*_*c*_, *ρ*_*t*_ measuring the correlation of the prognostic score with the outcome within each arm. Qualitatively, if the prognostic score strongly correlates with the outcome, the PPI estimator has a smaller variance than the standard one. If the prognostic score strongly negatively correlates with the outcome, one can reverse its sign and obtain the same result. However, a larger variance is obtained when the prognostic score weakly correlates with the outcome. This last case is highly undesirable as we do not want to risk losing power in a clinical trial setting.

### 2.3 Empowered estimators

#### 2.3.1 PPI++ estimator

In the extension of the initial PPI paper [2], the authors proposed circumventing the PPI method’s limitations by introducing a weighted contribution of the predictions and optimizing. The new estimator resulting from this operation is called the PPI++ estimator:

##### Definition 4

(PPI++ estimator). *Let λ* ∈ ℝ, *then:*

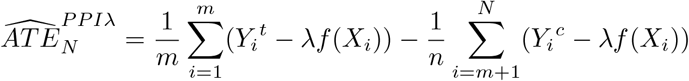

Notice that 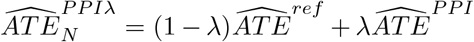. Therefore, this estimator is unbiased. Again, the CLT provides the following:

##### Proposition 3.

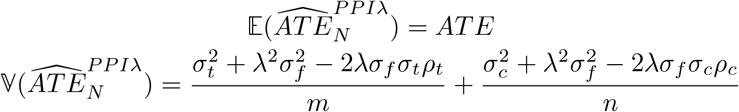

*and the estimator is asymptotically normal:*

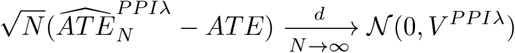

*where* 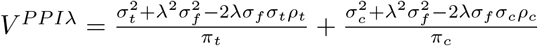

#### 2.3.2 PPCT estimator

We now define the Prediction-Powered for Clinical Trial (PPCT) estimator. Since *λ* is a design parameter, we can optimize it. We choose to minimize *V*^*PPIλ*^, which is equivalent to maximizing the power of the statistical test of the clinical trial. This fact will be discussed in section 2.4.2.

##### Definition 5

(PPCT estimator). *Let λ*\* *be the value minimizing the variance of the PPI++ estimator. The PPCT estimator is then:*

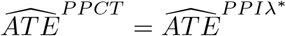

This definition can be made further explicit:

##### Proposition 4

(Optimal *λ*\* and variance of PPCT). *The PPCT estimator is unbiased for the ATE and:*

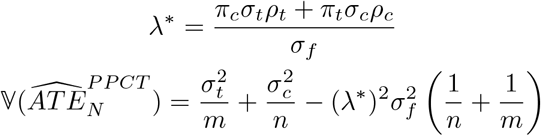

*with the asymptotical variance:*

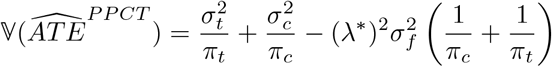

Comparing with Proposition 1, we obtain:

##### Corollary 5

(PPCT efficiency). *The PPCT estimator is always more efficient than the reference estimator for ATE*.

When computing the estimator, we use consistent estimators of the standard deviations and correlation coefficients to derive a consistent estimator of *λ*\*. In the following applications, we will use the empirical standard deviation and correlation computed from the data. The optimal value of *λ* indicates the quality of the predictor as well: the closer *λ*\* is to 1, the better the predictions. From a theoretical perspective, *λ*\* = 0 corresponds to the situation where *ρ*_*c*_ = *ρ*_*t*_ = 0, so that the predictor is uncorrelated with the outcome. In this case, the PPCT estimator is equal to the reference estimator. On the other hand, *λ*\* = 1 corresponds to *ρ*_*c*_ = *ρ*_*t*_ = 1. The prognostic score perfectly correlates with the outcome, and the PPCT estimator equals the PPI one.

Solving explicitly for an optimal partition of controls and treated patients is also an option: 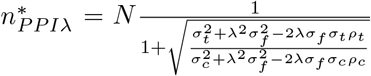. This expression can roughly be interpreted as follows: if the prognostic score correlates better with the outcome in the control arm than in the treated arm, then we increase the proportion of treated patients (and conversely). However, when designing the protocol, it is challenging to obtain reliable estimates of the prognostic score correlation with the outcome under no treatment *ρ*_*c*_ and, more especially, the correlation with the outcome after intervention *ρ*_*t*_, hence limiting the potential application of our method to the design of unbalanced arms trials.

##### 2.3.3 Parallel with prognostic covariate adjustment

We refer to the paper by [13] for more details about the formulas of prognostic covariate adjustment, which has received approval from both FDA and EMA [14] as a method to reduce clinical trial sample size. This method performs an ANCOVA using the prognostic score as a covariate.

###### Definition 6

(ANCOVA estimator). *Let us introduce the design matrix* **Z**^*T*^ = [**1, T**, *f* (**X**), **T***f* (**X**)]. *The outcome is modeled as a linear function of the design matrix:*

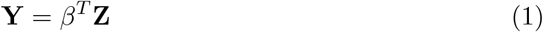

*where* 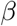 *are the coefficients of the regression and are estimated through ordinary least squares:* 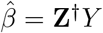. *The ANCOVA estimator of* 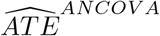, *corresponds to the second coordinate of* 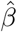.

The asymptotic variance of this estimator provided by [13] is:

###### Proposition 6.

*Assuming samples* (*Y*_*i*_, *X*_*i*_, *T*_*i*_)_*i*_ *are i*.*i*.*d, then:*

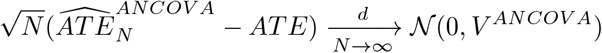

*where in the scalar case*, 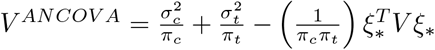 *with ξ*\* = *π* _*c*_ *ρ* _t_ *σ*_*t*_ *σ* _*f*_ + *π*_*t*_*ρ*_*c*_*σ*_*c*_*σ*_*f*_ *and V* = 𝕍 (*f* (*X*)).

This variance is the same as 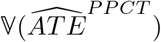. This should not come as a surprise as both are linear estimators adjusted with a prognostic score with the aim of minimizing variance. If we actually solve explicitly (1) in the case where dim(*f* (**X**)) = 1 (proof in supplementary), we obtain:

###### Proposition 7

(Unidimensional ANCOVA estimator). *If* dim(*f* (***X***)) = 1, *then:*

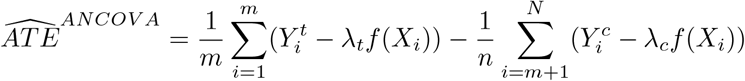

*where* 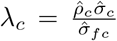 *and* 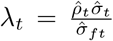, *with* 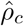 *being the empirical correlation between Y and* 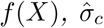 *the empirical standard deviation of Y and* 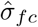 *the empirical standard deviation of f* (*X*) *under T* = 0 *(similarly superscript t applies with T* = 1).

This expression has many similarities to the 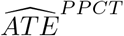 estimator, which instead of two different coefficients *λ*_*c*_ and *λ*_*t*_ has only one *λ*\*. However, being both consistent linear estimators, PPCT and ANCOVA are asymptotically equivalent, both in their pointwise ATE estimator and their variance estimator.

One noteworthy difference is that the variance of the PPCT estimator for a sample of size *N* has an analytic expression as a function of the model parameters, providing a more explicit and, thus, possibly more explainable estimator compared to ANCOVA. Another advantage of the PPI++ formulation is the possibility to generalize it to a non-linear setting, e.g., time-to-event data, see section 2.5.

### 2.4 Clinical Trial enhancement

#### 2.4.1 Statistical testing

Using the asymptotically normal distributions of the aforementioned estimators, we can perform two-sided tests for the null hypothesis *H*_0_: *ATE* = 0 vs the alternative *H*_0_: *ATE* ≠ 0. Namely:

##### Proposition 8.

*Let α* ∈ (0, 1) *be the desired type I error rate of the test. The null hypothesis is rejected when*

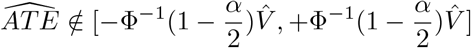

*where* Φ *is the cumulative distribution of the normal distribution and 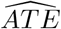 is an ATE estimator among* 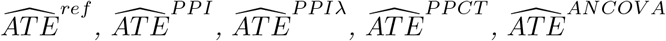 *and* 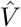 *is a consistent estimator of its asymptotic variance*.

As for standard clinical trial design, the normality assumption is valid only for large sample sizes, which is often considered to be the case when *n, m* ≥ 20. An alternative, valid for all the estimators described, is to use a permutation test [20] for assessing the success of the trial.

#### 2.4.2 Power and sample size computation

To compute the statistical test’s power, we introduce the treatment effect Δ_*H*_ under the alternative hypothesis.

##### Definition 7

(Power). *Let β the power of the statistical test in proposition 8*:

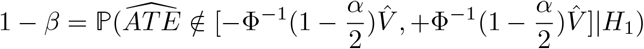

Under normality assumptions, the formula linking the power *β* of a trial to the sample size based on the hypothesized treatment effect size Δ_*H*_ and type-I error rate *α* is, therefore:

##### Proposition 9.

*Let α, β* ∈ (0, 1) *and* Δ_*H*_ ∈ ℝ. *The minimum sample size required for the statistical test in proposition 8 to have a level of α and a power of* 1 − *β is:*

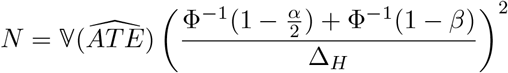

This shows how variance reduction directly affects the required sample size, with a proportional effect. A similar analysis would show the same proportional effect for a one-sided test when we are interested in the power to show a positive effect of active treatment, not just a difference (that could be negative). We undergo the routinely made hypotheses: *σ*_*c*_ = *σ*_*t*_ and *ρ*_*c*_ = *ρ*_*t*_ = *ρ* (which holds for a constant treatment effect, for instance), we obtain the following relationship for asymptotic variances of the estimators:

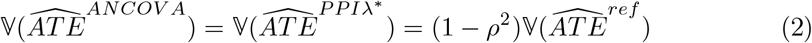

where *ρ*^2^ is actually *R*^2^, the out-of-sample R-squared.

This is particularly useful in clinical trial design as a rule to reduce sample size based on the estimated *R*^2^ of the prognostic factor. This formula implies that for the same power and type-I error control, the relative reduction in CT sample size is *R*^2^. We will verify whether this principle holds in our experiments on both simulated and real clinical trial data.

However, we want to emphasize that such simplifying assumptions are not likely to hold, especially *ρ*_*c*_ = *ρ*_*t*_, which assumes that the prognostic score explains as much variance of the treated arm as the control/placebo arm. This is very bold in the context of disease progression models, which are trained to predict the evolution under no intervention. We thus provide an analysis with more conservative assumptions.

We are interested in computing the power gain obtained using the PPCT estimator compared to the reference estimator as a function of the estimator’s performance. We focus here on reducing the size of the control arm.

Consider a standard design with a control and a treatment arm of the same size, notated m. The test statistic in proposition 8 uses 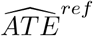. We are interested in comparing this design to an alternative one where the test statistic is the PPCT ATE estimator and the groups are unbalanced. The treatment group is of size *m*, the same as the standard design, while the control group is of size *n*\* ≤ *m*. We make the following assumptions:

1. We know *σ*_*c*_. We have access to a trained predictor *f*. We know *ρ*_*c*_. Without loss of generality, we assume that *ρ*_*c*_ *≥* 0. If this were not the case, we would replace *f* with −*f*;
2. *σ*_*t*_ = *σ*_*c*_ := *σ* This is a standard assumption in the design of clinical trials;
3. *ρ*_*t*_ ≥ 0. This expresses that the signs of *ρ*_*c*_ and *ρ*_*t*_ are the same. Note that this is a minimal but still restrictive condition.

We constrain the PPCT to have at least the same power as the reference estimator. This is obtained by having the variance of the PPCT estimator be less or equal to the variance of the reference estimator. This provides the following

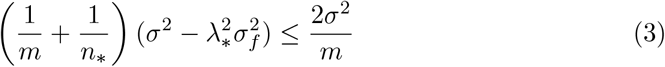

with

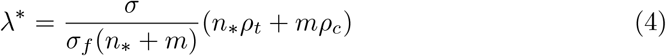

Simplifying, we obtain that

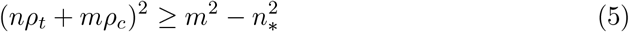

Using the fact that *ρ*_*c*_ *≥* 0 and *ρ*_*t*_ *≥* 0, we obtain that a sufficient condition is

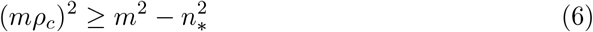

or equivalently,

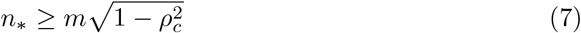

Thus, we can choose

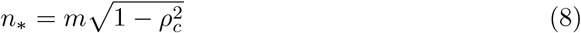

and obtain a design with the same power as the standard design. Note the following from (8)

1. when *ρ*_*c*_ = 0, *n*\* = *m* and the PPCT design is the same as the standard design;
2. when *ρ*_*c*_ = 1, *n*\* = 0, there is no need for a control group.

A range of values for *n*\**/m* are provided as a function of *ρ*_*c*_ in Table 1. A prediction highly correlated with the outcome (*ρ*_*c*_ large) would allow for a drastic reduction in the relative size of the control group compared to a standard balanced RCT

**Table 1.** Ratio of the size of the control group (*n*\*) to the size of the treatment group (*m*) for obtaining the same power as a standard balanced RCT. This ratio is shown as a function of the correlation between the classifier and the outcome (*ρ*_*c*_).

| $\rho_c$ | 0.0 | 0.1 | 0.2 | 0.3 | 0.4 | 0.5 | 0.6 | 0.7 | 0.8 | 0.9 | 1.0 |
| --- | --- | --- | --- | --- | --- | --- | --- | --- | --- | --- | --- |
| $\frac{n_*}{m}$ | 1.00 | 0.99 | 0.98 | 0.95 | 0.92 | 0.87 | 0.80 | 0.71 | 0.60 | 0.44 | 0.00 |

### 2.5 General prediction-powered for clinical trial estimator

The PPI method can be generalized to other estimators of interest in the context of clinical trials. We define the largest possible application framework, as in PPI++ [2], by focusing on a class of estimators resulting from the maximization of an objective function. Equivalently, such an estimator can be defined using a loss function *ℓ*_*θ*_ : (*X, Y*) ∈ 𝒳 × 𝒴 ↦ *ℓ*_*θ*_(*X, Y*) ∈ (*R*) parametrized by *θ* ∈ Θ, from which we define the population loss *L*(*θ*) = 𝔼 [*ℓ*_*θ*_(*X, Y*)], and the prediction loss *L*^*f*^ (*θ*) = 𝔼 [*ℓ*_*θ*_(*X, f* (*X*))]. The estimand is 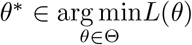. We introduce the relations to the data observations with the empirical losses:

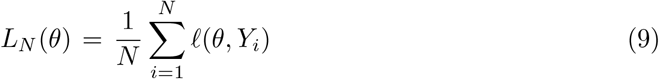

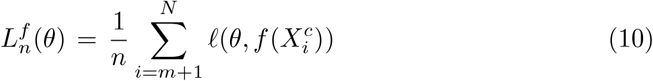

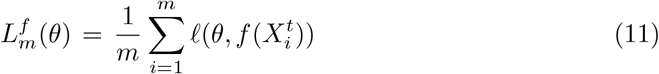

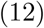

From there we define the PPCT loss:

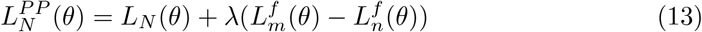

with *λ* ∈ ℝ. Using the fact that *X ⊥ T*, we have 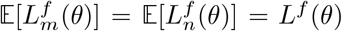, thus 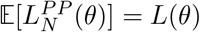. Then:

#### Definition 8

(General PPCT estimator). *Let the general PPCT estimator:*

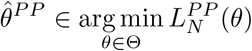

The PPI++ analysis [2] can be generalized to show that under some regularity assumptions, 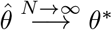 strongly. Many estimators fall under this generic framework, including linear regression (for instance, adjusted for covariates in an ANCOVA), logistic regression, and Cox regression. Details will be provided elsewhere.

## 3 Experiments and results

We will showcase the performance of the PPCT estimator on both synthetic data and real-life clinical trial data. Our application is focused on Alzheimer’s disease (AD) but is by no means restricted to AD or neurodegenerative diseases. The interest lies in the length of such chronic diseases, where the follow-up of patients has allowed the development of disease progression models. Such models have shown promising results for predicting the future evolution of the patients, for instance, in the TADPOLE challenge [21].

### 3.1 Simulations in unbalanced settings

In this first experiment, we present a simple simulation setting to illustrate the differences between the estimators under study. We particularly examine the performance of treatment effect estimators under unbalanced arms settings by varying the number of control patients and investigating how the quality of the prognostic score affects both the pointwise ATE estimators and their variance estimators. Let us describe the simulation model :

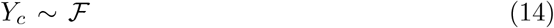

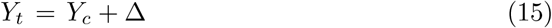

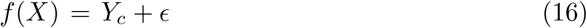

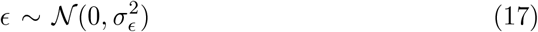

where *ϵ* is the noise of the predictor, with 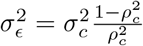. Notice that *ρ* = *ρ* since we consider a constant treatment effect. We fix the treatment effect Δ = 1. We evaluate two distributions, first the Normal (or Gaussian) distribution 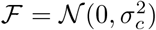 with *σ*_*c*_ = 1 and second the Chi-square distribution with 5 degrees of freedom to have a skewed distribution. We set the number of treated patients to *m* = 100 while varying the number of controls *n* = 2, 5, 10, 50, 100 to investigate how a strong predictor can facilitate the reduction in the number of control patients in a clinical trial. The results over 1000 simulations in each setting are shown in the tables 2 and C1 in the supplementary material.

**Table 2.** Mean ATE estimation ± one standard deviation estimated with 1,000 Monte-Carlo simulations. **Bold**: estimators with the lowest standard deviation per row. Red: estimators that are problematic, being either very biased or having an overly high variance.

| $\rho$ | n | Reference | PPI | PPCT | PPCT2 | ANCOVA |
| --- | --- | --- | --- | --- | --- | --- |
| **Normal** |  |  |  |  |  |  |
| 0.00 | 2 | **0.99 ± 0.71** | 0.99 ± 1.0 | 0.98 ± 0.95 | **0.99 ± 0.71** | **0.45 ± 20** |
|  | 5 | **1.0 ± 0.48** | 1.0 ± 0.67 | 1.0 ± 0.53 | **1.0 ± 0.48** | 1.0 ± 0.57 |
|  | 10 | **0.99 ± 0.32** | 0.99 ± 0.47 | 0.99 ± 0.33 | **0.99 ± 0.32** | 0.99 ± 0.34 |
|  | 50 | **1.0 ± 0.17** | 1.0 ± 0.25 | 1.0 ± 0.18 | 1.0 ± 0.18 | 1.0 ± 0.18 |
|  | 100 | **1.0 ± 0.14** | 1.0 ± 0.21 | **1.0 ± 0.14** | **1.0 ± 0.14** | **1.0 ± 0.14** |
| 0.50 | 2 | 0.99 ± 0.72 | 1.0 ± 1.2 | 1.0 ± 0.89 | **1.0 ± 0.62** | **1.1 ± 5.2** |
|  | 5 | 0.99 ± 0.46 | 1.0 ± 0.81 | 1.0 ± 0.45 | **1.0 ± 0.41** | 1.0 ± 0.47 |
|  | 10 | 1.0 ± 0.34 | 1.0 ± 0.58 | 1.0 ± 0.30 | **1.0 ± 0.29** | 1.0 ± 0.30 |
|  | 50 | 1.0 ± 0.18 | 0.99 ± 0.30 | **1.0 ± 0.15** | **1.0 ± 0.15** | **1.0 ± 0.15** |
|  | 100 | 1.0 ± 0.14 | 0.99 ± 0.24 | **0.99 ± 0.13** | **0.99 ± 0.13** | **0.99 ± 0.13** |
| 0.95 | 2 | **0.99 ± 0.73** | 1.0 ± 1.0 | 0.98 ± 0.98 | 1.0 ± 1.0 | **-0.34 ± 32** |
|  | 5 | 1.0 ± 0.46 | 1.0 ± 0.46 | **0.99 ± 0.41** | 1.0 ± 0.46 | 1.0 ± 0.45 |
|  | 10 | **0.99 ± 0.33** | 1.0 ± 0.38 | **1.0 ± 0.33** | **1.0 ± 0.33** | **1.0 ± 0.33** |
|  | 50 | 1.0 ± 0.17 | **0.99 ± 0.15** | **0.99 ± 0.15** | **1.0 ± 0.15** | **1.0 ± 0.15** |
|  | 100 | 1.0 ± 0.14 | **1.0 ± 0.13** | **1.0 ± 0.13** | **1.0 ± 0.13** | **1.0 ± 0.13** |
| **Chi-square** |  |  |  |  |  |  |
| 0.00 | 2 | **1.0 ± 2.2** | 1.1 ± 2.3 | **0.87 ± 3.2** | **1.0 ± 2.2** | **-2.6 ± 67** |
|  | 5 | **0.95 ± 1.5** | **0.96 ± 1.5** | 0.94 ± 1.6 | **0.95 ± 1.5** | 0.96 ± 1.7 |
|  | 10 | **0.99 ± 1.0** | 1.0 ± 1.1 | 1.0 ± 1.1 | **0.99 ± 1.0** | 1.0 ± 1.1 |
|  | 50 | **1.0 ± 0.55** | 1.0 ± 0.58 | 1.0 ± 0.56 | **1.0 ± 0.55** | 1.0 ± 0.56 |
|  | 100 | **0.98 ± 0.45** | 0.99 ± 0.48 | **0.98 ± 0.45** | **0.98 ± 0.45** | **0.98 ± 0.45** |
| 0.50 | 2 | 0.98 ± 2.3 | 0.98 ± 1.2 | **1.6 ± 2.1** | **0.96 ± 1.1** | **2.8 ± 52** |
|  | 5 | 0.98 ± 1.4 | 1.0 ± 0.79 | 1.3 ± 0.85 | **1.0 ± 0.69** | 1.1 ± 0.78 |
|  | 10 | 0.97 ± 1.1 | 1.0 ± 0.59 | 1.2 ± 0.61 | **1.0 ± 0.52** | 1.1 ± 0.55 |
|  | 50 | 1.0 ± 0.54 | 1.0 ± 0.30 | **1.0 ± 0.26** | **1.0 ± 0.26** | **1.0 ± 0.26** |
|  | 100 | 1.0 ± 0.45 | 1.0 ± 0.24 | **1.0 ± 0.21** | ±1.0 ± 0.21 | **1.0 ± 0.21** |
| 0.95 | 2 | 0.93 ± 2.3 | **1.0 ± 0.24** | **1.7 ± 2.1** | 0.99 ± 0.25 | **1.2 ± 7.3** |
|  | 5 | 0.96 ± 1.5 | **1.0 ± 0.15** | **1.4 ± 0.87** | 0.99 ± 0.16 | 1.0 ± 0.21 |
|  | 10 | 0.99 ± 1.0 | **1.0 ± 0.11** | **1.2 ± 0.33** | **1.0 ± 0.11** | 1.0 ± 0.12 |
|  | 50 | 0.99 ± 0.57 | **1.0 ± 0.06** | 1.0 ± 0.07 | **1.0 ± 0.06** | **1.0 ± 0.06** |
|  | 100 | 1.0 ± 0.47 | **1.0 ± 0.05** | **1.0 ± 0.05** | **1.0 ± 0.05** | **1.0 ± 0.05** |

Table 2 shows the values of five point-wise treatment effect estimators 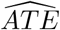: the reference, the PPI, the PPCT, and a variant denoted PPCT2, and the ANCOVA estimators. These estimators are described in the method section. The PPCT estimator uses 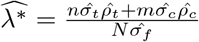 with 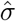 being the empirical standard deviation (with one degree of freedom to be unbiased) and 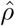 being the empirical correlation. This yields a consistent estimator for *λ*\*. However, note that 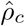 is being multiplied by *m* while 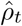 mis multiplied by *n*, which means that in extremely unbalanced settings with *n << m*, the variance of this estimator is high due to the dominant term being 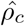 which is estimated over the few *n* subjects. Therefore, we proposed a variant, denoted PPCT2, for *λ*\*, which relies on a constant treatment effect assumption such that *ρ*_*c*_ = *ρ*_*t*_, i.e., the covariance of the predictor with the outcome is the same under treatment or without it. In this case, 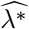 is estimated with 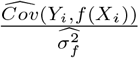 where the numerator is the unbiased pooled covariance estimator and the denominator is the standard unbiased estimator of the variance.

Table 2 highlights in red the estimators that are problematic, being either highly biased or exhibiting an overly high variance. It is the case systematically for ANCOVA for *n* = 2, that is, for a control group of size two, which is an extreme example. PPCT is also problematic for *n* = 2 when the response is Chi-squared distributed with 5 degrees of freedom. The Reference, PPI, and PPCT2 are always stable.

Excluding the case *n* = 2, we observe that under normally distributed data, none of the estimators appear to be biased. Then, in the case where the predictor is weakly correlated with the outcome (*ρ* ≤ 0.5), the PPCT estimators are competitive, and more specifically, the PPCT2 estimator almost always provides the least variable estimator.

In the case of balanced arms and even at *n* = 50, ANCOVA, PPCT and PPCT2 perform well and quite similarly.

With a skewed distribution, here the Chi-square with 5 degrees of freedom, we can observe deviations from the targeted *ATE* = 1 value at low control group sample sizes (*n* ≤ 5) for the PPCT estimator. However, PPCT2 fixes this issue and consistently outperforms the PPCT and ANCOVA. Lastly, in a highly correlated scenario (*ρ* = 0.95), the PPI estimator performs the best. However, this estimator is relatively poor if the predictor is less correlated with the outcome.

When using the Normal approximation of the test statistic, the pointwise ATE estimators are not enough to decide whether the treatment effect is significant or not; we also require the variance of these estimators. We use the following variance estimators: for the reference 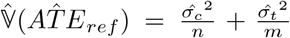, for the PPCT and PPCT2 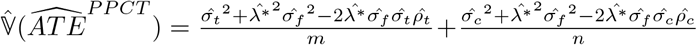 and we use the sandwich estimator for the least square regression of the ANCOVA. The results for the variance estimators are shown in Table C1 in the supplementaries.

From the results in the normal setting, we gather that low sample sizes in the control arm, i.e., *n* = 2, 5, tend to lead to a relatively high variance for the ANCOVA estimator. We also notice that in those extreme cases, the PPCT estimator tends to largely underestimate its true variance, which would lead to a high false discovery rate (FDR). Correcting with a better *λ*\* estimator as in PPCT2 yields an unbiased variance estimator. The PPCT2 variance estimator also consistently has lower variance than the PPCT and ANCOVA at lower sample sizes. At large sample sizes, however, PPCT, PPCT2, and ANCOVA have similar variance estimators, which match the best possible estimators (i.e., reference in the random predictor case *ρ* = 0 and PPI in the quasi-perfect predictor case *ρ* = 0.95).

Those results suggest that for balanced designs, PPCT and ANCOVA behave similarly. However, for designs enabled by the advent of prognostic models, the settings with low samples in the control arm are interesting. Here, some differences between the estimators appear. Namely, we see that PPCT would lead to a high FDR, while ANCOVA requires at least five patients in the control arm to be unbiased. However, the choice of estimator for *λ*\* in PPCT is of importance as showcased by PPCT2, which performs the best in terms of variance estimation at low *n* values while remaining unbiased. However, remember that the experimental design is here limited to *σ*_*c*_ = *σ*_*t*_.

### 3.2 Simulated clinical trial data

In those simulation experiments, we used the adsim R package [22]. The model used in this library [23] has been calibrated on AD patient data and provides close to reallife clinical trial setting simulations. The outcome of interest is the ADAS-Cog-13 for AD Assessment Scale-cognitive, a clinical score between 0 and 70, increasing with cognitive decline. The generative model for this score is Beta distributed:

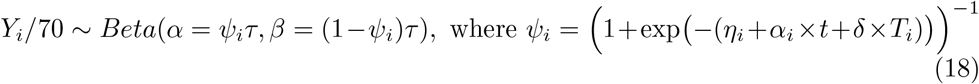

where *η*_*i*_ and *α*_*i*_ are random effects with normal distributions (generated by latent covariates), *t* is the length of the trial, *δ* is the effect of the treatment on the logit of the ADAS-Cog-13, *ψ*_*i*_ is the estimated underlying progression model for the ADAS-Cog-13, and *τ* is the precision of the Beta noise added to the model to generate the observations. In the following experiments, *t* is fixed at 1 year, *τ* and the hidden fixed effects are set at the mean values available in the adsim package. We use a 50*/*50 randomization between the treated and placebo arms. The parameters are fixed to be as close as possible to real life, and small perturbations did not lead to meaningful changes in the experiments. A sample of trajectories, both under the control and treatment arm is shown on figure 2 with the parameters used in the simulation and described below.

**Fig. 1.**
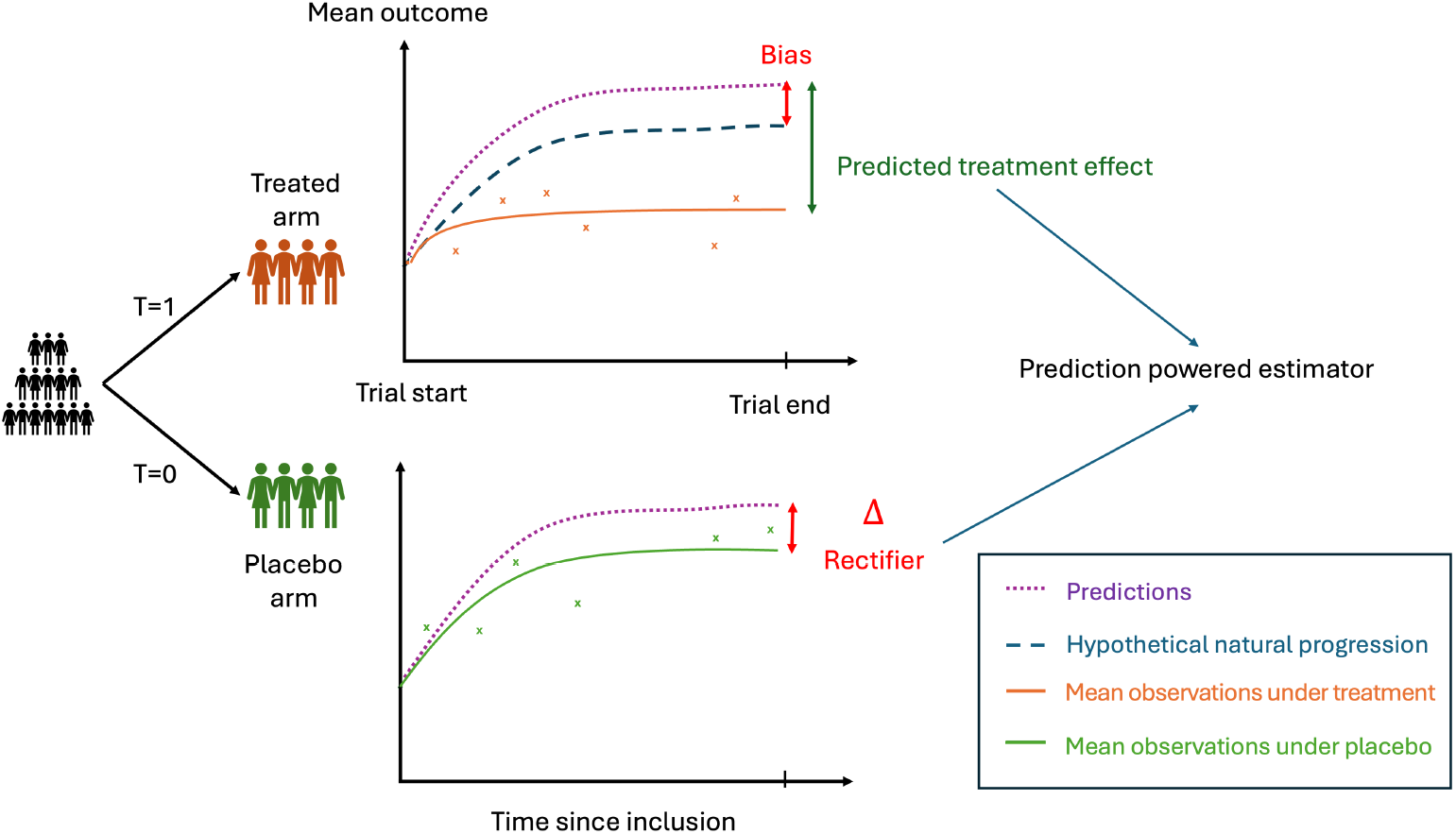
General scheme of the prediction-powered Inference estimator for Average Treatment Effect in a randomized clinical trial. The estimator leverages a prognostic model predicting the natural disease history of a patient using only information available at baseline. The estimator combines two terms: the predicted treatment effect, which is the difference between the observed outcome of treated patients and their predicted natural progression, and the rectifier, which accounts for the bias of the predictions (including both model error and placebo effect). The result is a new unbiased estimator whose variance depends on the quality of the predictions.

**Fig. 2.**
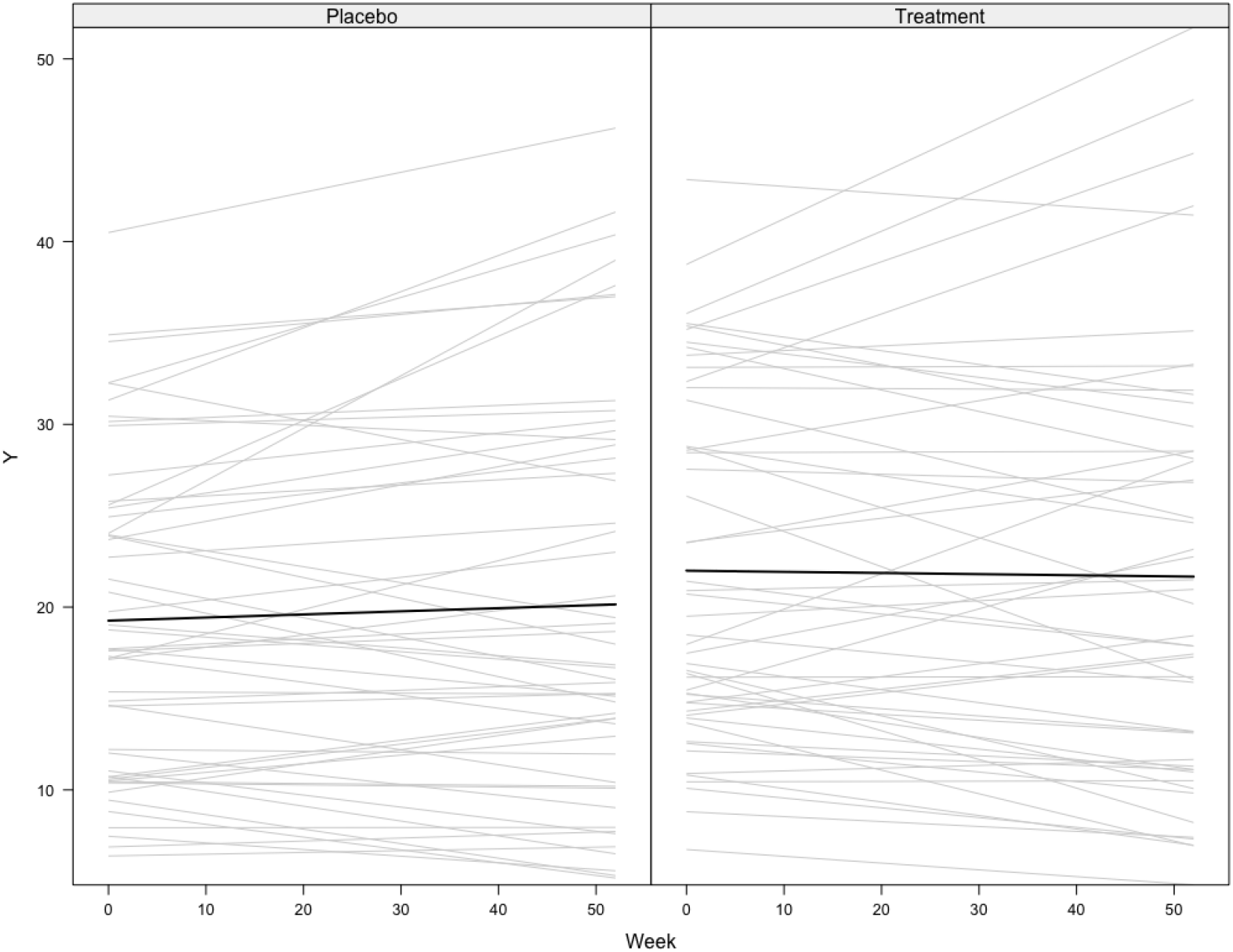
Trajectories of the ADAS-Cog-13 score progression, simulated for a sample of 100 placebo and treated patients across time, with a treatment size of *δ* = 0.2.

The only variables in the simulation model that we vary are the total number of subjects *N* and the precision of the Beta-distributed noise on the outcome *τ*.

To simulate a predictor, for each individual *i*, we use the underlying natural model *f* (*X*_*i*_) = *ψ*_*i*_(*η*_*i*_, *α*_*i*_; *T*_*i*_ = 0) + *ϵ*_*i*_, where *ϵ*_*i*_ ∼ 𝒩 (0, *σ*_*ϵ*_) allows us to control the predictor’s performance. Adding a constant bias did not influence the results since both PPI/PPCT and ANCOVA are adjusted linearly, so they account for any systematic bias.

We first simulated trials with *N* = 100 patients, with a treatment size being the one calibrated on the Donepezil in Alzheimer’s [23], *δ* = 0.1382 which is equivalent to an average treatment effect of minus two points over one year on the outcome scale: *ATE* ≃ −1.96. We simulated 1000 trials and gradually increased *σ*_*ϵ*_ to make predictions more noisy. We computed the power as the mean number of trials where the null hypothesis was rightly rejected. We also derived the theoretical sample size required for a trial equally powered as the standard design, i.e., using the reference estimator. The results are presented in Figure 3.

**Fig. 3.**
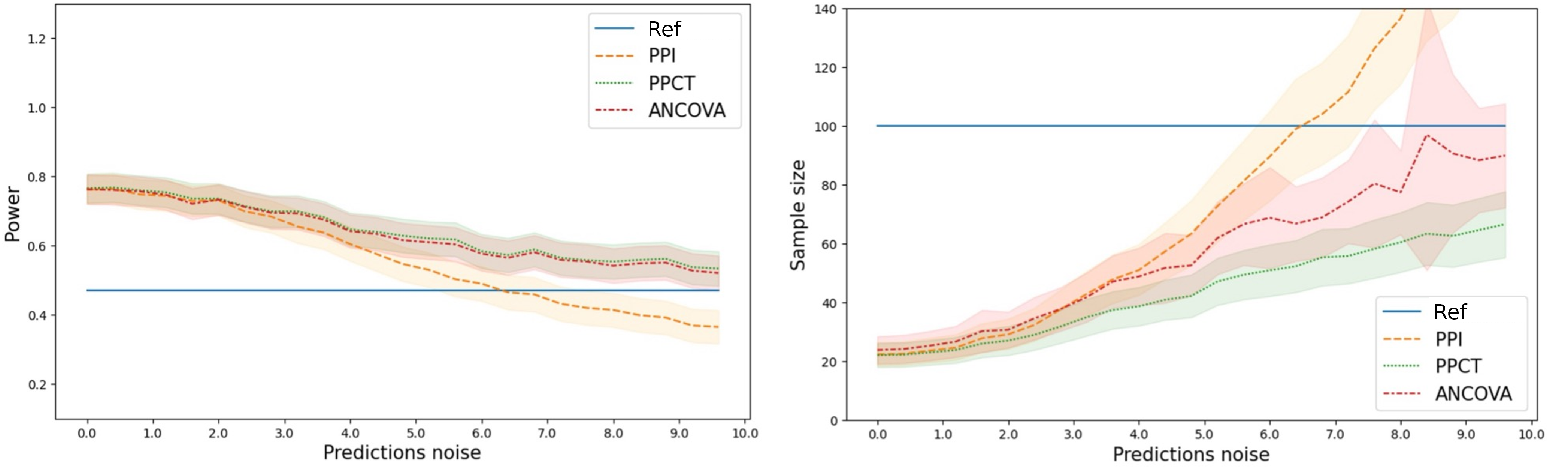
Left: Power of the estimators as a function of the prediction noise. Right: Sample size required for a clinical trial based on the different estimators as a function of the noise of the predictions. Noise is added on the native scale of the outcome (0-70). When the prediction noise is lower resp. higher than about 6.0, the PPI estimator is better resp. worse than the reference estimator. In contrast, PPCT and ANCOVA are always as or more powerful as the reference estimator. The reduction in sample size is more pronounced when the prediction is better (lower prediction noise).

We observe a gain of power for the empowered estimators PPCT and ANCOVA, which equates to a smaller sample size for the clinical trial. When the predictor is too noisy, the PPI estimator performs worse than the standard design. The PPCT and ANCOVA estimators are never significantly different, indicating their highly similar performance. They are always preferred to the standard design for this metric. The power increase and the sample size reduction are more pronounced when the predictions correlate more with the outcome. All these phenomena are consistent with the theory presented in the previous sections.

The PPCT with 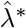 and ANCOVA estimators are consistent but not necessarily unbiased for a finite sample. Indeed, for PPCT, note that *λ*\* is estimated with the trial data. We provide an additional simulation experiment to evaluate the bias of the estimators. We varied the size of the clinical trial simulations, with a fixed noise level of *σ*_*ϵ*_ = 2.0 for the predictions. Figure 4 shows boxplots for all 4 methods at the sample sizes *N* = 10, 100, 200, 1000. In all cases, the bias is negligible.

**Fig. 4.**
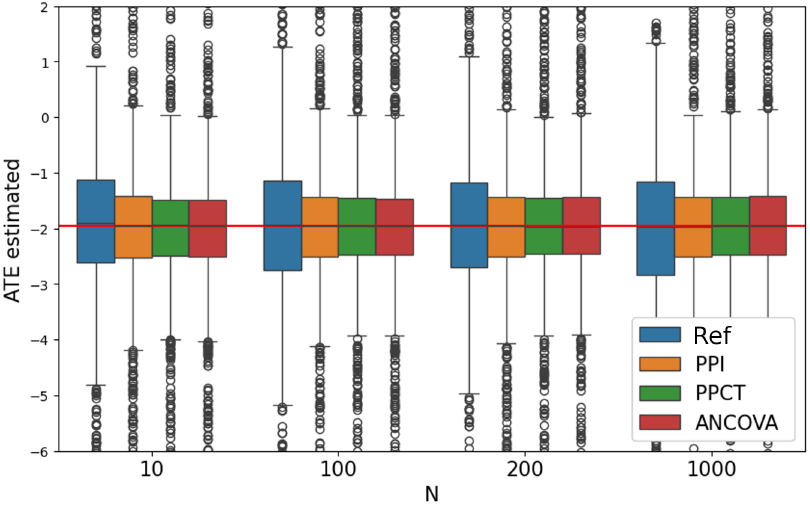
Average Treatment Effect estimated by 4 methods as a function of the trial size over 1000 simulations. Each method provides negligible bias. PPI, PPCT, and ANCOVA show a reduced variance and thus improve upon the reference estimator.

Simulations showcase the two main applications of the PPCT estimator: increase the power of a clinical trial or reduce the sample size at the design stage. Experiments with close to real-life data show that the estimated treatment effect has no bias.

### 3.3 Application to clinical trial data

#### 3.3.1 Dataset overview

We now apply the estimators to a real-life AD clinical trial: the Hippocampus Study [24]. This one-year clinical trial showed that Donepezil significantly reduced the annualized rate of atrophy in the hippocampus. Even though the primary endpoint of the trial was the atrophy of the hippocampus measured by MRI and automatically segmented by a pre-defined pipeline (Freesurfer [25]), the trial also recorded secondary outcomes, among which the volume of the ventricles and two clinical assessments: the Mini-Mental State Examination (a clinical assessment of cognitive abilities) and the ADAS13 (which was also the outcome in the previous simulations). After data processing, the values were available for *N* = 167 subjects, with *n* = 89 placebo and *m* = 78 treated patients. A descriptive summary of the variables can be found in Table 3.

**Table 3.** Summary of the Hippocampus Study values at baseline and end of trial. MMSE: Mini-Mental State Examination, cognitive assessment ranging from 0 (worse) to 30 (healthy); ADAS13: AD Assessment Scale, cognitive score ranging from 0 (healthy) to 70 (worse); MRI: Magnetic Resonance Imagery; ICV: intracranial volume, i.e. the ratio of the volume of the considered region (hippocampus or ventricles) over the total brain volume.

|  | MMSE |  | ADAS13 |  | MRI Hippocampus ICV |  | MRI Ventricles ICV |  |
| --- | --- | --- | --- | --- | --- | --- | --- | --- |
|  | Baseline | End | Baseline | End | Baseline | End | Baseline | End |
| count | 167 | 167 | 167 | 167 | 167 | 167 | 167 | 167 |
| mean | 26.0 | 25.3 | 20.2 | 21.8 | 0.004 | 0.004 | 0.026 | 0.027 |
| std | 3.1 | 3.0 | 5.1 | 6.4 | 0.001 | 0.001 | 0.010 | 0.011 |
| min | 0 | 17 | 9 | 7 | 0.003 | 0.003 | 0.006 | 0.006 |
| 25% | 25 | 23.5 | 16 | 18 | 0.004 | 0.004 | 0.019 | 0.020 |
| 50% | 27 | 26 | 20 | 22 | 0.004 | 0.004 | 0.024 | 0.026 |
| 75% | 28 | 27 | 24 | 25 | 0.005 | 0.005 | 0.030 | 0.033 |
| max | 30 | 30 | 35 | 47 | 0.006 | 0.006 | 0.067 | 0.071 |

#### 3.3.2 Predictors

For the prognostic scores, we used Disease Progression Models (DPM) calibrated on a natural disease history observational cohort.

##### Dataset

We used the Alzheimers Disease Neuroimaging Initiative (ADNI) database. This longitudinal cohort records a wide range of modalities, from MRI to biomarkers to clinical assessments. Namely, all the markers from the Hippocampus Study are included in this database, meaning that we can learn the typical progression patterns of those features. We will then be able to predict the progression of the trial’s patients using a well-calibrated DPM. The data’s descriptive statistics can be found in Table 4. We included all patients with an AD diagnosis and at least two visits, which boils down to 730 subjects for a total of 4636 visits.

**Table 4.** Summary of the ADNI variables. MMSE: Mini-Mental State Examination, cognitive assessment ranging from 0 (worse) to 30 (healthy); ADAS13: AD Assessment Scale, cognitive score ranging from 0 (healthy) to 70 (worse); MRI: Magnetic Resonance Imagery; ICV: intracranial volume, i.e. the ratio of the volume of the considered region (hippocampus or ventricles) over the total brain volume.

|  | MMSE | ADAS13 | MRI Hippocampus ICV | MRI Ventricles ICV |
| --- | --- | --- | --- | --- |
| count | 3799 | 3623 | 3395 | 3392 |
| mean | 23.789 | 27.018 | 0.004 | 0.032 |
| std | 4.706 | 11.816 | 0.001 | 0.014 |
| min | 0 | 0 | 0.001 | 0.005 |
| 25% | 22 | 19 | 0.003 | 0.022 |
| 50% | 25 | 26 | 0.004 | 0.030 |
| 75% | 27 | 33.330 | 0.004 | 0.040 |
| max | 30 | 85 | 0.007 | 0.090 |

##### Disease Progression Models

We calibrated three different models to predict the joint progression of the four selected markers (volume of hippocampus, volume of ventricles, MMSE and ADAS13):

1. Disease Course Mapping [7]: a non-linear mixed effect model based on Riemannian geometry. This model assumes a common pattern of progression in the population, determined by the population parameters, and that individuals deviate from it according to random effects captured by their parameters (for instance, an acceleration factor to distinguish between fast and slow progressors). We used the logistic curves Disease Course Mapping model, as it is well-suited for cognitive scores, such as the MMSE and ADAS13. The implementation of the model has been done with the Python library Leaspy
2. ODE-augmented-Lagrangian [8] is an algorithm that uses the augmented Lagrangian method of optimization for learning a non-parametric ordinary differential equation (ODE) of the form 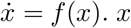 is a vector of biomarkers – assuming all the biomarkers are real-valued, 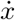 is the time-derivative of this vector, and *f* is a non-parametric function belonging to a reproducing kernel Hilbert space.
3. ODE-OCK [9] is the same model as for the ODE-augmented-Lagrangian optimized using the occupation kernel (OCK) formulation.

We then compute individualized predictions for the patients of the Hippocampus Study, using their values available at baseline and deducing the evolution at one year. However, this is a challenging task, as we only have one observation (at baseline), even though it is 4-dimensional, which is usually not enough to infer the underlying rate of progression. The prediction at one year is therefore less accurate than if we had visits for the patients prior to inclusion in the trial.

##### Results

We considered all four markers as potential trial endpoints. The outcome value *Y*_*i*_ for a patient *i* was obtained as the difference between their value at the end of the trial and their value at baseline. As the theoretical study shows, the predictor works best when the prognostic factor is highly correlated with the outcome. The prognostic factors were therefore adjusted to be the predicted change in score over one year, meaning that we took the difference between the predictions of the DPM and the value of the patient at baseline.

We computed the estimators and their variance, and deduced the required sample size to have an equally powered clinical trial using formula provided in Proposition 9. The results are shown in table 5.

**Table 5.** Sample size required for an equally powered clinical trial using the estimators with three predicting models on the four endpoints of the Hippocampus Study. DCM: Disease Course Mapping model; OCK: occupation kernel model; Lagrangian: augmented Lagrangian ODE model. MMSE: Mini-Mental State Examination; ADAS13: AD Assessment Scale; MRI: Magnetic Resonance Imagery. Boldface highlights the minimum of each column when it is less than 100

|  | MMSE | ADAS13 | MRI Hippocampus | MRI Ventricles |
| --- | --- | --- | --- | --- |
| Reference | 100 | 100 | 100 | 100 |
| PPI (DCM) | 94 | 172 | 572 | 2639 |
| PPCT (DCM) | 84 | **97** | 100 | 100 |
| ANCOVA (DCM) | **76** | 99 | 103 | 103 |
| PPI (OCK) | 133 | 187 | 597 | 2307 |
| PPCT (OCK) | 100 | 100 | 100 | **88** |
| ANCOVA (OCK) | 103 | 101 | 103 | 91 |
| PPI (Lagrangian) | 127 | 204 | 599 | 2565 |
| PPCT (Lagrangian) | 100 | 98 | 100 | 100 |
| ANCOVA (Lagrangian) | 107 | 99 | 104 | 103 |

We observe a sample size reduction for the PPCT and ANCOVA estimators in the case of the MMSE and ADAS13 scores, as well as the MRI Ventricles. The best-performing models are the DCM for the MMSE and ADAS13 and the OCK for the MRI ventricles. The DCM model reaches an *R*^2^ of 15% on the MMSE. This number means that we can either increase the power of the trial or reduce the sample size to obtain an equally powered trial. The DCM predictions’ correlation is better if we only look at controls, *ρ*_*c*_ = 0.625, whereas it does not capture as well the evolution of treated patients, *ρ*_*t*_ = 0.125. This is coherent with the purpose of a DPM, as it accurately predicts the progression of placebo patients, while the model is not meant to describe the trajectories of patients under treatment.

The OCK method shows a sample size reduction on the ventricles, with an *R*^2^ around 10%, *ρ*_*c*_ = 0.337 and *ρ*_*t*_ = 0.375. In practice, the correlation of the predictions with the outcome is greatly limited by the noise inherent to the measure. For the MRI measures, since the variability of the measure far outweighs the scale of the changes over one year, the *R*^2^ of the predictions with the outcome is thus capped. The *R*^2^ obtained did not differ much from 0 on the hippocampus, even with the models based on ODEs, which are less constrained than the DCM and have an easier time learning the progression of the imagery features.

From a statistical test standpoint, the reduction of the variance did not lead to a significant enough confidence interval reduction to change the conclusions of the trial on the modification of the MMSE nor the ADAS13, as seen in Figure 5. On the specific instance of the MMSE, we observe that the reduction in the confidence interval is counterbalanced by the estimator being closer to zero. For the hippocampus, the PPCT and ANCOVA do not change the estimator, thereby leading to the same conclusion as the primary analysis of the trial. The reduction of variance on the ventricles brought the estimator closer to statistical significance. We can imagine that with an improved prediction, for instance, if we had more points per patient prior to the baseline, the test could have rejected the null hypothesis.

**Fig. 5.**
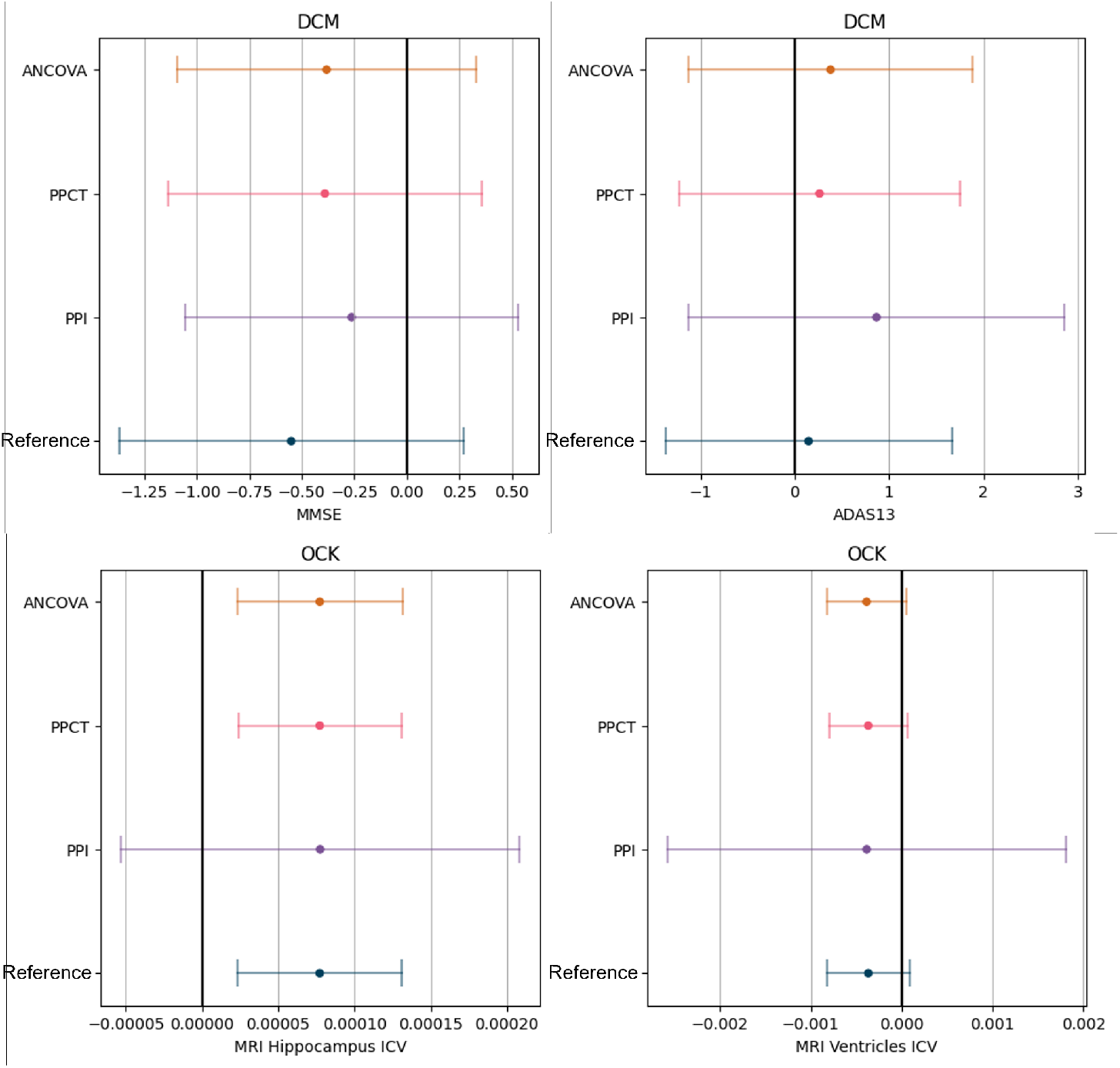
Estimators with their 95% confidence interval on the 4 outcomes of the Hippocampus Study. The predictor used for the prediction-powered estimators and the ANCOVA is the Disease Course Mapping model for the two cognitive scores (MMSE and ADAS13) whereas we showed the results with the occupation kernel ODE method for the imagery markers (hippocampus and ventricles).

## 4 Discussion

The newly introduced prediction-powered estimator for Clinical Trials (PPCT) is a powerful tool to enhance clinical trials. This method leverages historical data through a prognostic score and refines the estimator for the Average Treatment Effect. We have demonstrated that it is unbiased and decreases the variance of the estimator compared to the reference method. These properties have been validated on simulated clinical trial data. Our method has mainly two applications: either increasing the power of the clinical trial while maintaining the same type I error rate, or reducing the sample size at the design stage while retaining an equally powered RCT.

Our theoretical study shows that if we fix the tuning hyperparameter *λ*, the estimator is unbiased even at small sample sizes and has a known variance. When estimating this parameter using the trial data, the bias is shown empirically to be negligible. If we use assumption-free tests such as a permutation test, we do not require the estimator’s asymptotic distribution to decide the trial’s result. This can be particularly interesting when the trial size cannot guarantee the normality assumption. The estimator is very similar to the prognostic covariate adjustment method at large sample sizes. We derived the explicit formulation of the latter to show the differences, even if they are minor.

One point that needs to be addressed is whether the parameter *λ* should be fixed in advance of the statistical analysis, based on prior knowledge of the outcome and prognostic score correlations (from a pilot study or historical data), or computed “on the fly” with a consistent estimator of the optimal *λ* as we did in our experiments. This allows for a better estimator but loosens the theoretical guarantees about unbiasedness. Our simulations show that the bias is negligible. However, this point needs further exploration. Other methods to determine the parameter *λ*\* might be better and should be investigated in the future. As shown in the simulations, different estimators of the *λ*\* parameters lead to very different results. For instance, the direct consistent estimator leads to underestimation of the variance in low sample sizes, which could lead to a high false discovery rate. On the other hand, a more adequate *λ*\* estimator yields a much better *ATE* estimator.

We also provided an analysis of the variance estimators of the different ATE estimators, which are essential to determining whether the trial showed a significant effect. Under large sample sizes, we showed that the variance of the estimators is all consistent, with the variance of the PPCT estimator being dependent on the choice of 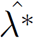. In the best case, the variance of the PPCT variance estimator can be lower than that of ANCOVA, which could lead to a higher power.

We chose to focus on the specific estimator for the Average Treatment Effect with a continuous outcome, as it encompasses the larger part of the RCT. However, we briefly introduced the general PPCT estimator, which is much more powerful. Further developments will allow us to apply this method to other critical estimators, for instance M-estimators, Cox regression, or even being able to adjust for covariates in the ATE estimation. This shows great promise for generalisation, as opposed to prognostic covariate adjustment, which can only be used in linearly adjusted estimation of the treatment effect on a continuous outcome.

The performance of the predictions is crucial, as a prognostic score highly correlated with the outcome can lead to large sample size reductions. Even without making too strong assumptions, we are able to derive sample size formulas to help decrease the sample size with accurate predictions. We showed that the prediction-powered framework favors unbalanced arms when the predictor is reliable. Indeed, one can interpret the predictions as synthetic control patients, thus lessening the need for actual controls in the trial. However, the method also performs sample size reduction in balanced trials, and this involves less risk at the design stage.

In our application to a real-life RCT, we obtained around 15–20% variance (or equivalently, sample size) reduction on secondary endpoints. The estimator of the treatment effect on the primary endpoint was not affected, as the prediction models were not able to forecast the outcome precisely at the individual level. This shows that the method can have practical implications, but also that it is hard to obtain strong prognostic factors. The challenge is complex, as we need a model able to predict the change in score of a patient over one year. This can be translated as: we need to capture the slope of each patient, not solely its intercept. Furthermore, the prognostic score needs to correlate with individual variability, meaning that imputing the mean slope is not enough. Doing so with only baseline covariates is a daunting task.

We can think of various ways to improve on the proposed predictors, for instance, using ensemble methods such as Super Learner [26]. Another route would be to increase the information available for each patient, namely past observations, in order to have more reliable predictions. It is easier to learn the slope from several points.

One perk of the technique is that the estimator and the prognostic factors can be engineered separately, allowing the trial designer to make use of any complex machine learning system built by another as a “black-box” predictor. This is all the more relevant as supervised learning techniques are developing rapidly, and are otherwise complicated to use in clinical practice. For instance, we can mention Deep Learning models, which are notoriously hard to interpret and are thus shunned by clinicians.

Our proposed PPCT estimator has desirable properties for a RCT: smaller variance, unbiasedness, and a simple explicit formula. The method can either be used to reduce the cost of a clinical trial by shrinking its size or in a safer way by increasing the power of the trial.

Overall, this paper proposes an application of the prediction-powered inference technique to clinical trials, considering the case of linear covariate adjustment. The derived estimators of the average treatment effect are shown to be different but closely related to existing ones, including the ANCOVA estimator, providing an alternative point of view, but also possible advantages in unbalanced trials and long-tailed outcomes. PPI and PPI++ were originally presented in the context of M-estimators. We foresee that bringing this research to the setting of clinical trials will provide benefits in non-linear settings, including qualitative and time-to-event outcomes.

## Data Availability

The data used in this article were obtained from the Alzheimers Disease Neuroimaging Initiative (ADNI) database (adni.loni.usc.edu), launched in 2003 as a public-private partnership, led by Principal Investigator Michael W. Weiner, MD. The primary goal of ADNI has been to test whether serial magnetic resonance imaging (MRI), positron emission tomography (PET), other biological markers, and clinical and neuropsychological assessment can be combined to measure the progression of mild cognitive impairment (MCI) and early Alzheimers disease (AD). The versions of the dataset used for this experiment were ADNI 1, 2, 3 and ADNI GO.
The data of the ‘Hippocampus Study’ were obtained thanks to Bruno Dubois and collaborators.

https://adni.loni.usc.edu

## 5 Declarations

### 5.1 Ethics approval and consent to participate

The ADNI data has been used in conformity with the ADNI terms of use detailed at https://adni.loni.usc.edu/terms-of-use/. The protocol of the ‘Hippocampus study’ and informed consent forms were approved by the ethics committee of Salpêtriere Hospital. The data has been used in conformity with the recommendations of this ethics committee.

### 5.2 Consent for publication

Not applicable

#### 5.3 Availability of data and materials

The data used in this article were obtained from the Alzheimers Disease Neuroimaging Initiative (ADNI) database (adni.loni.usc.edu), launched in 2003 as a public-private partnership, led by Principal Investigator Michael W. Weiner, MD. The primary goal of ADNI has been to test whether serial magnetic resonance imaging (MRI), positron emission tomography (PET), other biological markers, and clinical and neuropsychological assessment can be combined to measure the progression of mild cognitive impairment (MCI) and early Alzheimers disease (AD). The versions of the dataset used for this experiment were ADNI 1, 2, 3 and ADNI GO.

The data of the Hippocampus Study were obtained thanks to Bruno Dubois and collaborators and were first published in [24].

#### 5.5 Funding

The research leading to these results has received funding from the program “Investissements davenir” ANR-10-IAIHU-06. This work was also funded in part by the French government under management of Agence Nationale de la Recherche as part of the “Investissements d’avenir” program, reference ANR-19-P3IA-0001 (PRAIRIE 3IA Institute). The work at Portland State University was partly funded by the National Institute of Health RO1AG021155, R01EY032284, R01AG027161, and National Science Foundation #2136228. Data collection and sharing for this project was funded by the Alzheimer’s Disease Neuroimaging Initiative (ADNI) (National Institutes of Health Grant U01 AG024904) and DOD ADNI (Department of Defense award number W81XWH-12-2-0012). ADNI is funded by the National Institute on Aging, the National Institute of Biomedical Imaging and Bioengineering, and through generous contributions from the following: AbbVie, Alzheimers Association; Alzheimers Drug Discovery Foundation; Araclon Biotech; BioClinica, Inc.; Biogen; Bristol-Myers Squibb Company; CereSpir, Inc.; Cogstate; Eisai Inc.; Elan Pharmaceuticals, Inc.; Eli Lilly and Company; EuroImmun; F. Hoffmann-La Roche Ltd and its affiliated company Genentech, Inc.; Fujirebio; GE Healthcare; IXICO Ltd.; Janssen Alzheimer Immunotherapy Research & Development, LLC.; Johnson & Johnson Pharmaceutical Research & Development LLC.; Lumosity; Lundbeck; Merck & Co., Inc.; Meso Scale Diagnostics, LLC.; NeuroRx Research; Neurotrack Technologies; Novartis Pharmaceuticals Corporation; Pfizer Inc.; Piramal Imaging; Servier; Takeda Pharmaceutical Company; and Transition Therapeutics. The Canadian Institutes of Health Research is providing funds to support ADNI clinical sites in Canada. Private sector contributions are facilitated by the Foundation for the National Institutes of Health (www.fnih.org). The grantee organization is the Northern California Institute for Research and Education, and the study is coordinated by the Alzheimers Therapeutic Research Institute at the University of Southern California. ADNI data are disseminated by the Laboratory for Neuro Imaging at the University of Southern California.

#### 5.6 Authors’ contributions

An author contribution table is given in table 6.

**Table 6.** Contributions. PEP: Pierre-Emmanuel Poulet, MT: Maylis Tran, ST: Sophie Tezenas du Montcel, SD: Stanley Durrleman, BMJ: Bruno Michel Jedynak, BD: Bruno Dubois

| Contribution | PEP | MT | ST | SD | BMJ | BD |
| --- | --- | --- | --- | --- | --- | --- |
| Conception | ✓ |  |  | ✓ | ✓ |  |
| Design | ✓ |  |  | ✓ | ✓ |  |
| Acquisition |  |  |  |  |  | ✓ |
| Analysis | ✓ | ✓ |  | ✓ | ✓ |  |
| Interpretation | ✓ |  | ✓ | ✓ | ✓ |  |
| Software creation | ✓ |  |  |  |  |  |
| Draft or revised | ✓ | ✓ | ✓ | ✓ | ✓ | ✓ |

## 5.7 Acknowledgements

Not applicable

## Footnotes

* Data used in preparation of this article were obtained from the Alzheimers Disease Neuroimaging Initiative (ADNI) database (adni.loni.usc.edu). As such, the investigators within the ADNI contributed to the design and implementation of ADNI and/or provided data but did not participate in analysis or writing of this report. A complete listing of ADNI investigators can be found at: http://adni.loni.usc.edu/wp-content/uploads/howtoapply/ADNI Acknowledgement List.pdf

