## Supplementary Material for "Prediction-powered Inference for Clinical Trials: application to linear covariate adjustment"

#### Appendix A Explicit derivation of ANCOVA estimator

In our case, we compare the model where there is no baseline covariates, thus the design matrix is: $Z^T = [\mathbf{1}, \boldsymbol{T}, f(\mathbf{X}), \mathbf{T}f(\mathbf{X})]$. The system to solve is:

$$\boldsymbol{Y} = Z^T\boldsymbol{\beta} \tag{A1}$$

The least-square solution is explicitly given by: $\boldsymbol{\beta}^* = (ZZ^T)^{-1}Z\mathbf{Y}$. We have:

$$A = ZZ^T = \begin{bmatrix} N & m & \sum_i f(X_i) & \sum_i T_i f(X_i) \\ m & m & \sum_i T_i f(X_i) & \sum_i T_i f(X_i) \\ \sum_i f(X_i) & \sum_i T_i f(X_i) & \sum_i f(X_i)^2 & \sum_i T_i f(X_i)^2 \\ \sum_i T_i f(X_i) & \sum_i T_i f(X_i) & \sum_i T_i f(X_i)^2 & \sum_i T_i f(X_i)^2 \end{bmatrix} = \begin{bmatrix} B & C^T \\ C & D \end{bmatrix}$$

To invert this matrix we use the Schur complement : $S = D - CB^{-1}C^T$ which, after some calculus, is equal to :

$$\begin{aligned} S &= \begin{bmatrix} n\hat{\sigma}^2_{fc} + m\hat{\sigma}^2_{ft} & m\hat{\sigma}^2_{ft} \\ m\hat{\sigma}^2_{ft} & m\hat{\sigma}^2_{ft} \end{bmatrix} \\ \hat{\sigma}^2_{fc} &= \left(\frac{1}{n}\sum_i (1-T_i) f(X_i)^2\right) - \left(\frac{1}{n}\sum_i (1-T_i) f(X_i)\right)^2 \\ \hat{\sigma}^2_{ft} &= \left(\frac{1}{m}\sum_i T_i f(X_i)^2\right) - \left(\frac{1}{m}\sum_i T_i f(X_i)\right)^2 \end{aligned}$$

We then use the following formula to invert the matrix :

$$A^{-1} = \begin{bmatrix} B^{-1} + B^{-1}C^T S^{-1} C B^{-1} & -B^{-1}C^T S^{-1} \\ -S^{-1}CB^{-1} & S^{-1} \end{bmatrix}$$

We don't actually need to compute all the coefficients of the inverse, only the ones in the second row are enough to compute $\beta_2$, the coefficient associated with the treatment :

$$
\begin{aligned}
\beta_2 &= (A^{-1}ZY)_2 = A^{-1}_{2,:}ZY \\
&= \frac{1}{m}\sum_i T_i(Y_i - \frac{\hat{\rho_t}\hat{\sigma_t}}{\hat{\sigma_{ft}}}f(X_i)) - \frac{1}{n}\sum_i (1-T_i)(Y_i - \frac{\hat{\rho_c}\hat{\sigma_c}}{\hat{\sigma_{fc}}}f(X_i)) \\
\hat{\sigma_c} &= \left(\frac{1}{n}\sum_i (1-T_i)Y_i^2\right) - \left(\frac{1}{n}\sum_i (1-T_i)Y_i\right)^2 \\
\hat{\sigma}_t^2 &= \left(\frac{1}{m}\sum_i T_iY_i^2\right) - \left(\frac{1}{m}\sum_i T_iY_i\right)^2 \\
\hat{\rho_c} &= \frac{\hat{Cov}(f_c(X), Y_c)}{\hat{\sigma}_c\hat{\sigma}_{fc}} \\
\hat{Cov}(f_c(X), Y_c) &= \frac{1}{n}\sum_i (1-T_i)f(X_i)Y_i - \left(\frac{1}{n}\sum_i (1-T_i)f(X_i)\right)\left(\frac{1}{n}\sum_i (1-T_i)Y_i\right) \\
\hat{\rho_t} &= \frac{\hat{Cov}(f_t(X), Y_t)}{\hat{\sigma}_t\hat{\sigma}_{ft}} \\
\hat{Cov}(f_t(X), Y_t) &= \frac{1}{m}\sum_i T_if(X_i)Y_i - \left(\frac{1}{m}\sum_i T_if(X_i)\right)\left(\frac{1}{m}\sum_i T_iY_i\right)
\end{aligned}
$$

Therefore, if we rewrite it with our notations:

$$
\hat{ATE}^{ANCOVA} = \frac{1}{m}\sum_{i=1}^{m}(Y_{i|T_i=1} - \lambda_t f(X_i)) - \frac{1}{n}\sum_{i=m+1}^{N}(Y_{i|T_i=0} - \lambda_c f(X_i)) \tag{A2}
$$

where $\lambda_c = \frac{\hat{\rho_c}\hat{\sigma_c}}{\hat{\sigma_{fc}}}$ and $\lambda_t = \frac{\hat{\rho_t}\hat{\sigma_t}}{\hat{\sigma_{ft}}}$, to be compared with PPCT which instead of two different coefficients has: $\lambda^* = \frac{n\sigma_t\rho_t + m\sigma_c\rho_c}{\sigma_f N}$ which is the arithmetic mean of the two in expectation. $\forall a \in \{0,1\}, \hat{\rho_a} = \frac{\hat{Cov}(f(\mathbf{X}),Y|T=a)}{\hat{\sigma_a}\hat{\sigma_{fa}}}, \hat{Cov}(f(\mathbf{X}), Y|T = a) = \frac{\sum_i \mathbb{1}(T_i=a)(Y_i-\bar{Y}_a)(f(X_i)-\bar{f(X)}_a)}{\sum_i \mathbb{1}(T_i=a)}, \hat{\sigma_a}^2 = \frac{\sum_i \mathbb{1}(T_i=a)(Y_i-\bar{Y}_a)^2}{\sum_i \mathbb{1}(T_i=a)}, \hat{\sigma_{fa}}^2 = \frac{\sum_i \mathbb{1}(T_i=a)(f(X_i)-\bar{f(X)}_a)^2}{\sum_i \mathbb{1}(T_i=a)}, \bar{Y}_a = \frac{\sum_i \mathbb{1}(T_i=a)Y_i}{\sum_i \mathbb{1}(T_i=a)}, \bar{f(X)}_a = \frac{\sum_i \mathbb{1}(T_i=a)f(X_i)}{\sum_i \mathbb{1}(T_i=a)}$

### Appendix B  Consistent estimators nuances

As mentioned, in practice we use a consistent estimator for $\lambda^*$ in the PPCT estimator. This does not guarantee unbiasedness of our estimator anymore, therefore asking the validity of its use. In practical experiments we will show that this is negligible even at small sample sizes. If anything, the estimator is more conservative about the treatment effect size. Indeed, if we introduce $\hat{D}_N = A\hat{T}E_N^{standard} - A\hat{T}E_N^{PPCT}$, we have:

$$Cov(A\hat{T}E_N^{standard}, \hat{D}_N) = \lambda^* \left( \frac{1}{m} Cov(Y, f(X)|T=1) + \frac{1}{n} Cov(Y, f(X)|T=0) \right) \tag{B3}$$

where $Cov(Y, f(X)|T = 0)$ (resp. $T = 1$) is the covariance of the outcome $Y$ with the prognostic score $f(X)$ on placebo (resp. treated) patients. Using consistent estimators of the covariance, $C_{fY}^t = Cov(Y, f(X)|T = 1)$ (resp. $C_{fY}^c \hat{Cov}(Y, f(X)|T = 0)$), and variance of $f(X)$, $\hat{V}_f$, for $\lambda^*$ yields:

$$\hat{\lambda^*} = \frac{n\hat{C}_{fY}^t + m\hat{C}_{fY}^c}{N\hat{V}_f} \tag{B4}$$

$$\hat{Cov}(A\hat{T}E_N^{standard}, \hat{D}_N) = \frac{(n\hat{C}_{fY}^t + m\hat{C}_{fY}^c)^2}{nmN\hat{V}_f} \tag{B5}$$

Hence $\hat{Cov}(A\hat{T}E_N^{standard}, A\hat{T}E_N^{standard} - A\hat{T}E_N^{PPCT}) \geq 0$, so in practice the PPCT estimator tends to be more conservative.
